# Clinical features and outcomes of 197 adult discharged patients with COVID-19 in Yichang, Hubei

**DOI:** 10.1101/2020.03.26.20041426

**Authors:** Fating Zhou, Xiaogang Yu, Xiaowei Tong, Rong Zhang

**Affiliations:** The First College of Clinical Medical Science, China Three Gorges University, Yichang, Hubei, 443003, China; Emergency Department, Yichang Central People’s Hospital, Yichang, Hubei, 443003, China; Yichang third People’s Hospital, Yichang, Hubei, 443003, China

**Keywords:** Coronavirus infection, Acute respiratory distress syndrome, Pneumonia, COVID-19

## Abstract

**Purpose:** To investigate the epidemiology and clinical features of discharged adult patients with coronavirus infection disease 2019 (COVID-19) in Yichang.

**Method:** The retrospective study recruited 197 cases of COVID-19 discharged from Yichang Central People’s Hospital and Yichang Third People’s Hospital from Jan 17 to Feb 26, 2020. All cases were confirmed by real-time RT-PCR or chest computer tomography (CT). The survivors were followed up until March 4,2020. Clinical data, including demographic characteristic, presentation, underlying illness, exposure history, laboratory examination, radiology and prognosis were enrolled and analyzed by SPSS 19.0 software.

**Results:** There were 197 adult discharged patients with COVID-19 in this study. Statistical analysis indicated that the average age was 55.94 years, and female patients were 99(50.3%).Those patients mainly resided in urban with exposure history in 2 weeks, while 7 medical staffs were infected. Fever (77.6%%), cough (43.6%) and weakness (14.7%) were the common symptoms. Leukocytes were mainly normal or decreased in 185 patients (92.9%), both lymphocytes and eosinophils were below normal range, the ratios were 56.9% and 50.3%, respectively. On the contrary, lactate dehydrogenases raised in 65 patients. C-reactive protein (72.4%) elevated in the most of patients. The sensitivity of RT-PCR was 63.5%. Chest CT indicated that bilateral patchy shadows (69.0%) were the most common imaging manifestations.169(85.8%) patients recovered and transferred to a designated hospital for observation, and the others (14.2%) turned worst and died of acute respiratory failure.

**Conclusion:** COVID-19 infection with highly contagious have become a life-threaten public healthy problem, the sensitivity of RT-PCR was limited. Chest CT scan was recommended for the suspected patients. Furthermore, lymphocytopenia and eosinophils declining without leukocytes increasing may be considered as a useful evidence for the diagnosis.

## Introduction

Since several cases of 2019 novel coronavirus (2019-nCoV) pneumonia was reported in Wuhan^[1]^.This disease was identified by the Chinese Center for Disease Control and Prevention(CDC) On January 3, 2020. And then was named as COVID-19 by WHO ^[2]^. At the end of January, this disease spread rapidly to other cities of adjacent to Wuhan in Hubei province, including Huanggang, Xiaogan, Jingzhou and Yichang. Yichang, the 2^nd^ largest city with Three Gorges Dam in Hubei province, was about 300 kilometers far away from Wuhan, and many residents who worked or studied at Wuhan, returned to hometown for Spring Festival. By March 4, 2020, official reported there was 931 confirmed cases with COVID-19 in Yichang.

Similar to the server acute respiratory syndrome (SARS) and Middle East respiratory (MERS), 2019-nCoV can also cause serve acute respiratory syndrome ^[3].^ Numerous patients can be cured effectively at the early stage, but some patients with COVID-19 may develop pulmonary edema, ARDS, or multiple organ failure, and even died abruptly ^[4]^. Currently, researches about epidemiology and clinical features of COVID-19 focused on the hospitalized patients ^[5]^, but the discharged patients was still scarcely investigate. Therefore, this study was aimed to clarify the epidemiology and clinical features of discharged patients with COVID-19.

## Methods

This study was approved by the Ethics Committee of Yichang Central people’s hospital. All adult discharged patients with COVID-19 from Yichang Central People’s Hospital and Yichang third People’s Hospital from Jan 17 to Feb 26, 2020. Those patients with exposure histories were confirmed by chest CT scans or RT-PCR assays. Throat•swab or broncholveolr lvge fluid sample were collected from all the suspected patients at admission, and RT-PCR assays were performed at clinical laboratory. According to the 5th edition expert conference of Chinese healthy commission, those suspected patients presenting with lung imaging features also regard as clinically diagnosis-cases in Hubei province. ARDS was identified in accordance with the Berlin definition ^[6]^, and acute kidney injury was verified according to Improve Global Outcome definition ^[7]^. If the serum levels of cardiac biomarkers ascended to the 99th percentile upper reference limit or new abnormalities were displayed in echocardiography or electrocardiography (ECG), the patient was regarded as acute cardiac injury. The discharged criteria included that symptom disappeared and the lesions eliminated evidently in the chest CT. Furthermore, RT-PCR was negative continual 2 tests beyond 24 hours. After discharged from hospital, the patients went to the designated hospital for observation. All patients except deaths were monitored up to March 4, 2020.

### Statistical Analysis

Clinical data, including date, ages, occupation, location, presentation, underlying diseases, laboratory test, RT-PCR, chest CT, treatment and outcome was collected from electronic medical record(EMR). If data was missing, we collected by direct communication with attending doctor. Then the data was inputted and analyzed by SPSS 19.0 software. Descriptive statistics were presented as median with interquartile range or frequency with percentage.

## Results

### Epidemiology of COVID-19 patients in Yichang

A total of 197 adult discharged patients with COVID-19 were recruited in Yichang Central People’s Hospital(101, 51.3%)and Yichang third people’s hospital(86,48.7%).The first cases was a 65-years old woman from Wuhan on January15, 2020. The quantity of admitted patients increased sharply on the 15 days later, ascended to the peak on the February 2, and decreased obviously on Februray 7 (See the Figure1).

The average age of discharged patients was 55.94±18.83 (IQR, 40.25-70.75) years, and percentage of 49.7 (98) was female. Those cases were mainly occurred in the urban (178, 90.4%), especially Xiling District, because numerous patients were employees of China Gezhouba Group Corporation (CCGC) returning from Wuhan. Except for seven medical staffs, most of them were employees (58, 29.4%), retirees (55, 27.4%), and agriculture workers (29, 14.7%). A total of 187 patients were clustered and had clear exposure histories. Among of them, 75(38.7%) patients resided in the community of COVID-19 patients, 71(36.8%) patients had ever contacted with relatives or friends from Wuhan, 41(20.8%) patients had a trip to Wuhan.

### Clinical features of adult discharged patients with COVID-19

Fever (153, 77.6%) was the most common presentations, following cough (89, 45.2%) and weakness (29, 14.7%), respectively. Other less common symptoms were sore throat (11, 5.6%), shortness of breath (10, 5.1%), and muscle ache (9, 4.6%, Table 2). However, 3 patients without any complaints were detected by chest CT scan. Among all those patients, the average days from exposure to symptom onset was 6.14±9.27days. The longest incubation period was 55 days. 48 patients were accompanied with underlaying illness, including cardiovascular (24.4%) and cerebrovascular diseases (4.6%), endocrine system diseases (9.1%), chronic renal disease (1.5%), malignant tumor(1.5%), and digestive system diseases (1.0%), nervous system diseases. Considering vital sign and CT manifestations, 56 patients were admitted to intensive cure unite (ICU), and 141 patients were transferred to general isolation wards.

**Table 1.**
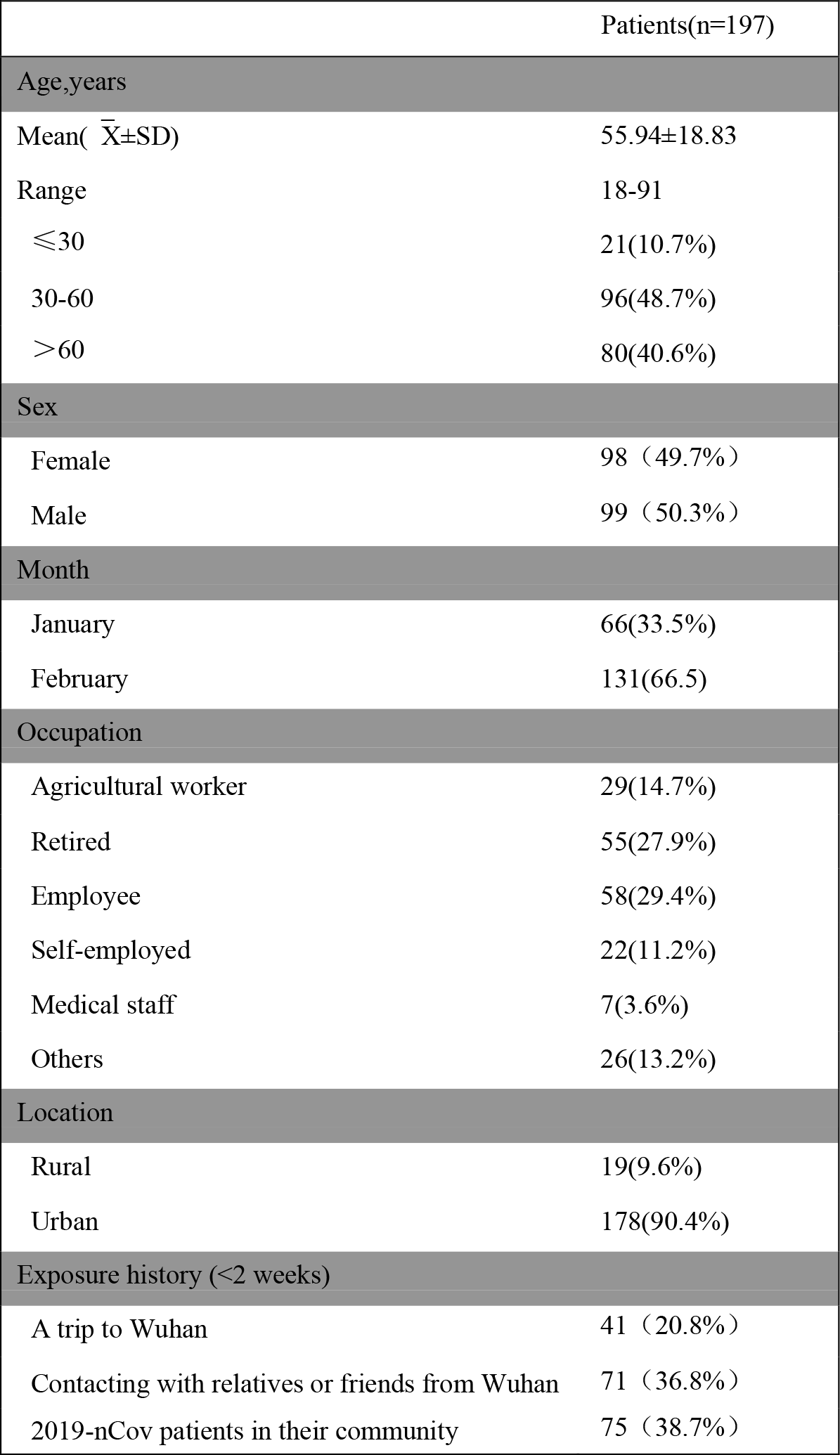
**Demographics and baseline characteristics of 197 discharged patients with COVID-19**.

**Table 2.**
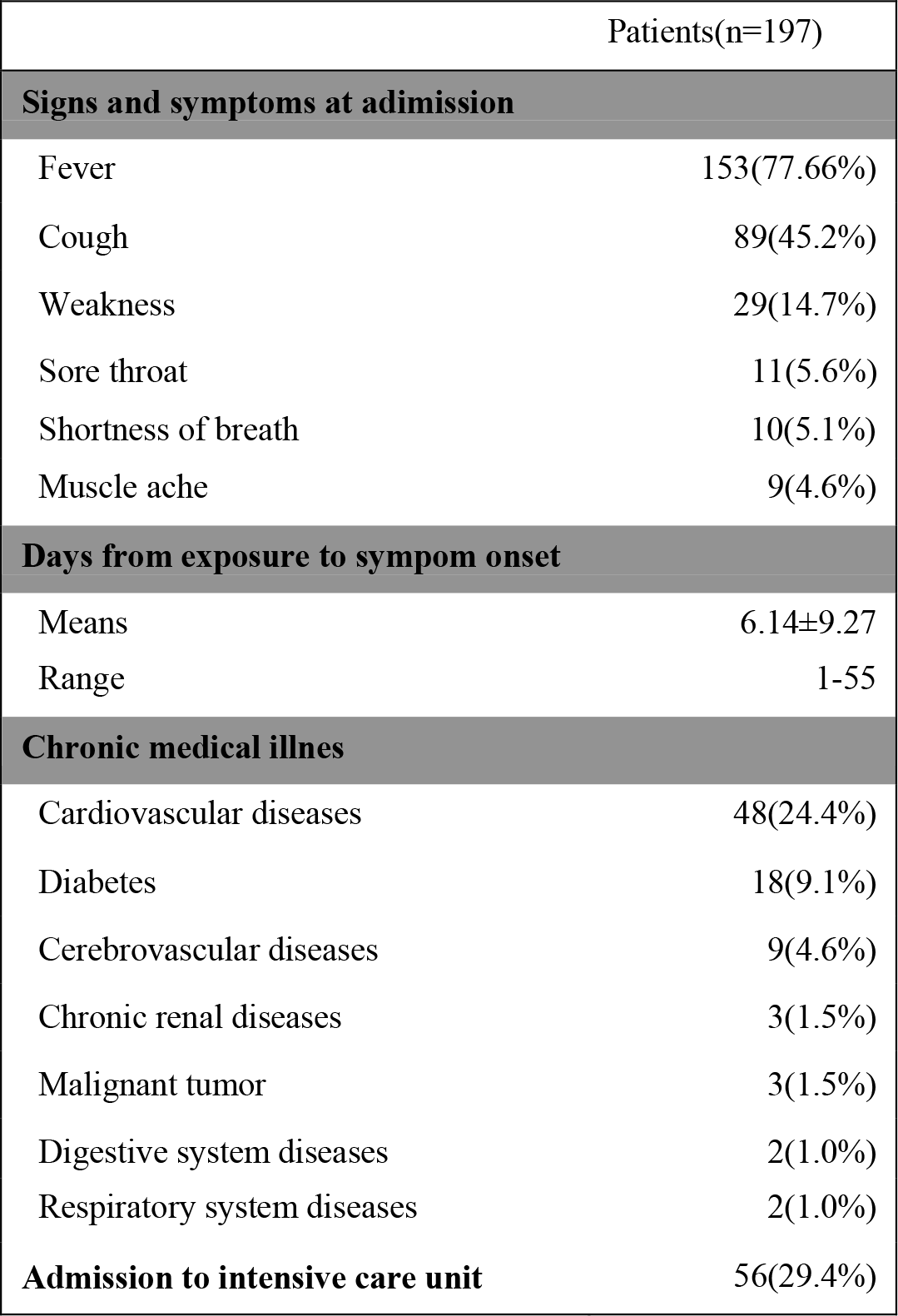
**Clinical characteristics of 197 discharged patients’ with COVID-19**.

**Table 3.** **Results of laboratory examinations in the 197 adult discharged patients with COVID-19**.

|  | Average | Normal Range | Increased | Decreased |
| --- | --- | --- | --- | --- |
| **Blood routine** |  |  |  |  |
| Leucocytes | 5.79±3.13 | 4-10 ($\times 10^9$ / L) | 14(7.1%) | 66(33.5%) |
| Neutrophils | 4.18±3.06 | 1.8-6.3 ($\times 10^9$ / L) | 33(16.8%) | 23(11.7%) |
| Eosinophils | 0.09±0.51 | 0.02-0.52 ($\times 10^9$ / L) | 1(5.1%) | 99(50.3%) |
| Lymphocytes | 1.13±0.74 | 1.1-3.2 ($\times 10^9$ / L) | 7 (3.6%) | 112 (56.9%) |
| Monocytes | 0.41±0.22 | 0.1-0.6 ($\times 10^9$ / L) | 4(2.0%) | 31(15.7%) |
| Platelet | 165.35±67.97 | 100-400 ($\times 10^9$ / L) | 1(0.5%) | 30(15.2%) |
| **Blood chemistry** |  |  |  |  |
| Alanine aminotransferase | 38.40±44.42 | 9-50 (U/ L) | 36(18.3%) | 7(35.5%) |
| Aspartate aminotransferase | 38.84±44.42 | 15-40 (U/ L) | 44(22.3%) | 11(5.6%) |
| Total bilirubin | 16.31±13.86 | 5.1-28 (μmol/L) | 24(12.2%) | 2(1.0%) |
| Direct bilirubin | 6.15±12.35 | 0-10.0 (μmol /L) | 10(5.1%) | – |
| Blood urea nitrogen | 7.69±9.86 | 3.1-8.0 (μmol /L) | 47(23.9%) | 52(26.4%) |
| Serum creatinine | 106.80±150.53 | 57-97 (μmol /L) | 35(17.8%) | 53(26.9%) |
| **Myocardial enzymes** |  |  |  |  |
| Creatine kinase | 133.67±223.48 | 50-310 (IU/L) | 10(5.1%) | 39(19.8%) |
| Creative kinase MB | 21.11±57.00 | 0-25 (IU/ L) | 19(9.6%) | – |
| Lactate dehydrogenase | 266.20±153.70 | 120-250 (IU/L) | 65(33.0%) | 8(4.1%) |
| Hydroxybutyrate dehydrogenase | 189.96±116.88 | 95-250 (IU /L) | 30(15.2%) | 14(7.1%) |
| **Coagulation function** |  |  |  |  |
| Prothrombin time | 11.18±1.72 | 10.0-14.5(s) | 6(3.0%) | 14(7.1%) |
| Activated partial thromboplastin time | 32.66±7.15 | 20.0-45.0(s) | 10(5.1%) | 2(1.0%) |
| Fibrinogen | 4.13±2.64 | 2.00-4.00(g /L) | 78(39.6%) | – |
| D-dimmer | 2309.36±8217.50 | 0-500(ng/ mL) | 93(47.2%) | – |
| **Blood gas analysis** |  |  |  |  |
| pH | 7.40±0.10 | 7.35-7.45 | 15/69(21.7%) | 11/69(15.9%) |
| PaO<sub>2</sub> | 81.77±37.73 | 80-100 (mmHg) | 4/69(5.8%) | 28/69(40.5%) |
| PaCO<sub>2</sub> | 37.73±9.53 | 35-45 (mmHg) | 8/69(11.6%) | 25/69(36.2%) |
| **Infection related biomarkers** |  |  |  |  |
| Procalcitonin | 0.46±2.17 | 0-0.05 (ng/ mL) | 60/176(34.1%) | – |
| C-reactive protein | 54.97±58.76 | 0-10 (mm /h) | 80/123(72.4%) | – |
| Erythrocyte sedimentation rate | 32.00±27.50 | 0-10 (mm/h) | 59/82(71.2%) | – |
| Interleukin-6 | 112.69±254.68 | 0-7 (pg/mL) | 23/35(65.7%) | – |

### Laboratory examinations of adult discharged patients with COVID-19

The laboratory examination was underwent after admission, the blood routine results revealed that monocytes and platelet kept at normal range in numerous patients, leukocytes were at the normal range in 117 (59.4%) patients, and below the normal range in 66 (33.5%) patients. While lymphocytes decreased in many patients. Interestingly, eosinophils displayed a similar change tendency as lymphocytes, it decreased obviously over half of them. 44 (22.3%) patients had different degrees of liver function abnormality, with elevating of aspartate aminotransferase (AST) or alanine aminotransferase (ALT). Total bilirubin (TBIL) was above the normal range in 24(12.2%) patients, and direct bilirubin (DBIL) was above in 10 (5.1%) patient. Most of those patients had a abnormal myocardial zymogram, which indicated that creatine kinase was above in 10 (5.1%) patients, creatine kinase MB in 19 (9.6%) patients, and lactate dehydrogenase in 65(33.0%) patients. For the coagulation tests, prothrombin time (PT) and activated partial thromboplastin time (APTT) were at the normal range in the majority of patients, while fibrinogen was above the normal range in the 78 (39.6%) patients. D-dimmer was above the normal range in the 99 (47.2%) patients. 65 (33.0%) patients had different degrees of renal function damage, with elevating blood urea nitrogen or creatinine. Blood gas analysis was examined in 69 cases, and the results demonstrated that PaO_2_ was decreased in the 28(40.5%) patients. Regarding the infection index, procalcitonin(PCT) mainly kept at the normal range. On the contrary, c-reative protein (CRP), erythrocyte sedimentation rate (ESR) and interleukin-6 (IL-6) increased in most of patients.

Although RT-PCR was regarded the most accurate examinations for COVID-19, the sensitivity was limited. RT-PCR positive rate was 63.5%, the first test was 46.7%, and the second test was 11.2%. Comparing with PCR, almost all patients had been discover lesions in the chest CT scans, 32 (15.3%) patients displayed unilateral pneumonia, 136 (69.0%)patients displayed bilateral pneumonia, and only 25(12.7%) patients displayed multiple mottling and ground-glass opacity(Table 4,Figure 2).

**Table 4.** **PCR, chest CT scans and treatment of discharged patients**.

|  |  |
| --- | --- |
| **Total positive of RT-PCR** | **63.5%** |
| The first test | 46.7% |
| The Second test | 11.2% |
| The third test | 4.1% |
| **Chest CT scan or X-ray** |  |
| Abnormities | 193(98.0%) |
| Unilateral pneumonia | 32(16.24%) |
| Bilateral pneumonia | 136(69.0%) |
| Multiple mottling and ground-glass opacity | 25 (12.7%) |
| **Treatment** |  |
| Oxygen therapy | 147(76.6%) |
| No-invasive | 34(15.3%) |
| invasive | 16(8.1%) |
| Antibiotic | 151(76.6%) |
| Antiviral | 179(90.9%) |
| Glucocorticoids | 75(38.1%) |
| Intravenous immunoglobulin | 52(26.9%) |
| Chinese Traditional medicine | 81(41.1%) |
| **Complication** |  |
| ARDS | 26(13.2%) |
| Acute renal injury | 16(8.1%) |
| Acute myocardial injury | 14(7.1%) |
| Septic shock | 12(6.1%) |
| Acute liver injury | 8(4.1%) |
| **Outcome** |  |
| Death | 28(14.2%) |
| Survival | 169(85.8%) |
| **Hospital stays (days)** |  |
| Means | 15.08±17.47 |
| 0-7 (days) | 79(40.1%) |
| 7-14 (days) | 31(15.7%) |
| >14 (days) | 87(44.2%) |

**Fig 1.**
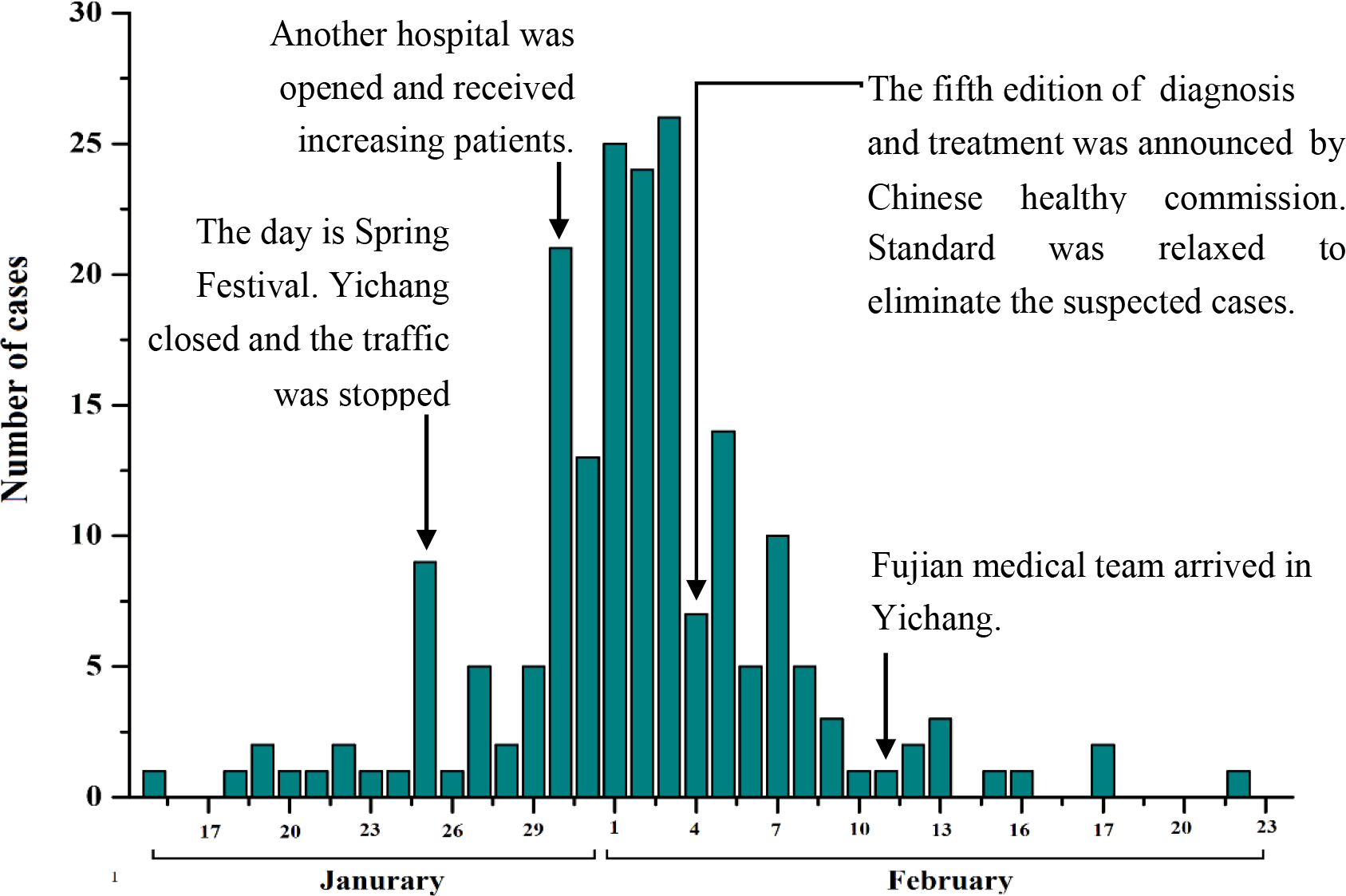
The distribution of admission date and crucial events in Yichang.

**Fig2.**
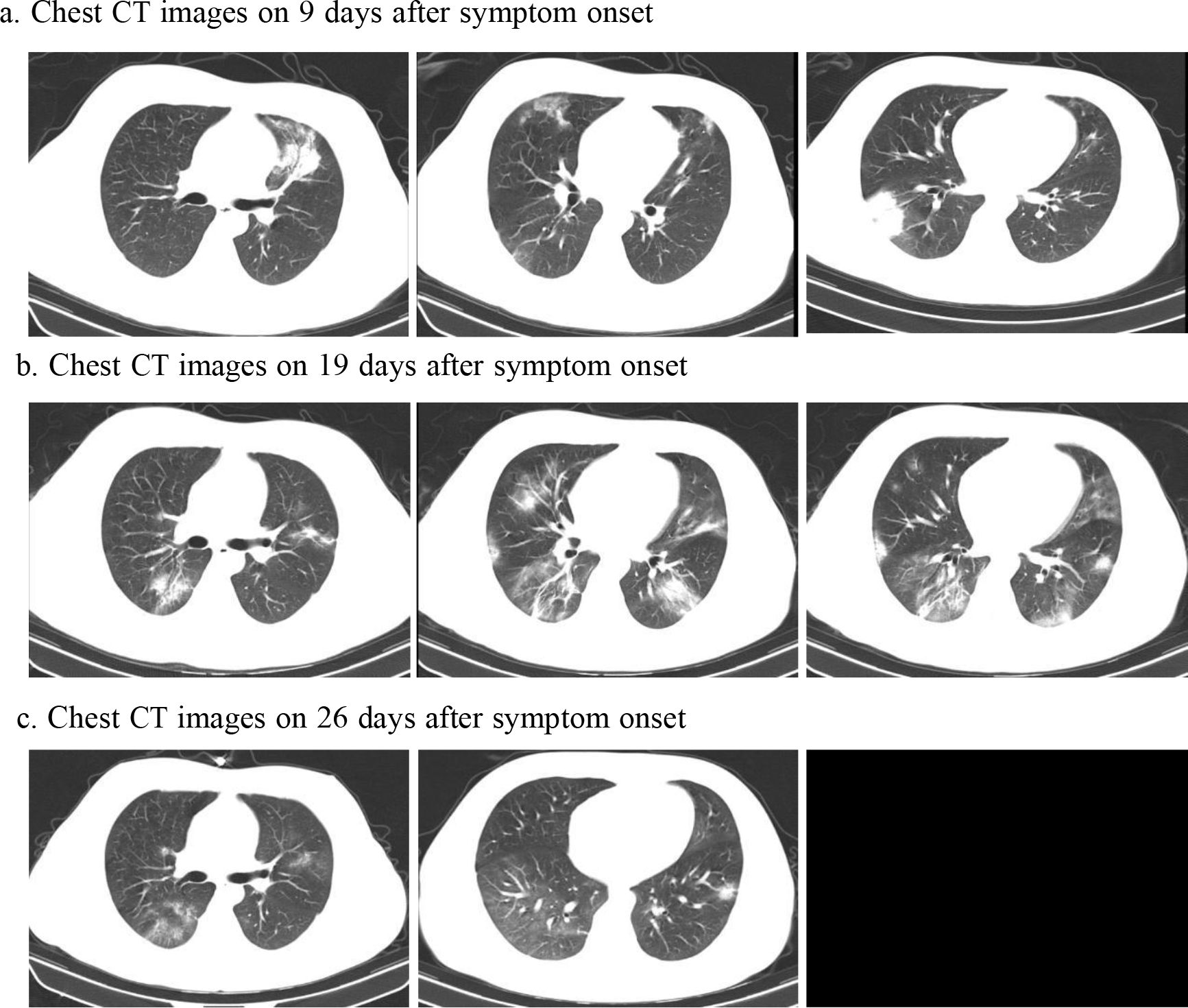
Chest CT images of a 26-year-old patient with COVID-19. a Chest CT images obtained on February 2, 2020, showed ground glass opacity in both lungs on day 5 after symptom onset. b Images taken on February 12,2020, indicated the lesions increased obviously. C Images taken on February 19, 2020.revealed bilateral ground glass opacity absorbed dramatically, and the patient discharged from hospital and return to the local hospital.

**Fig3.**
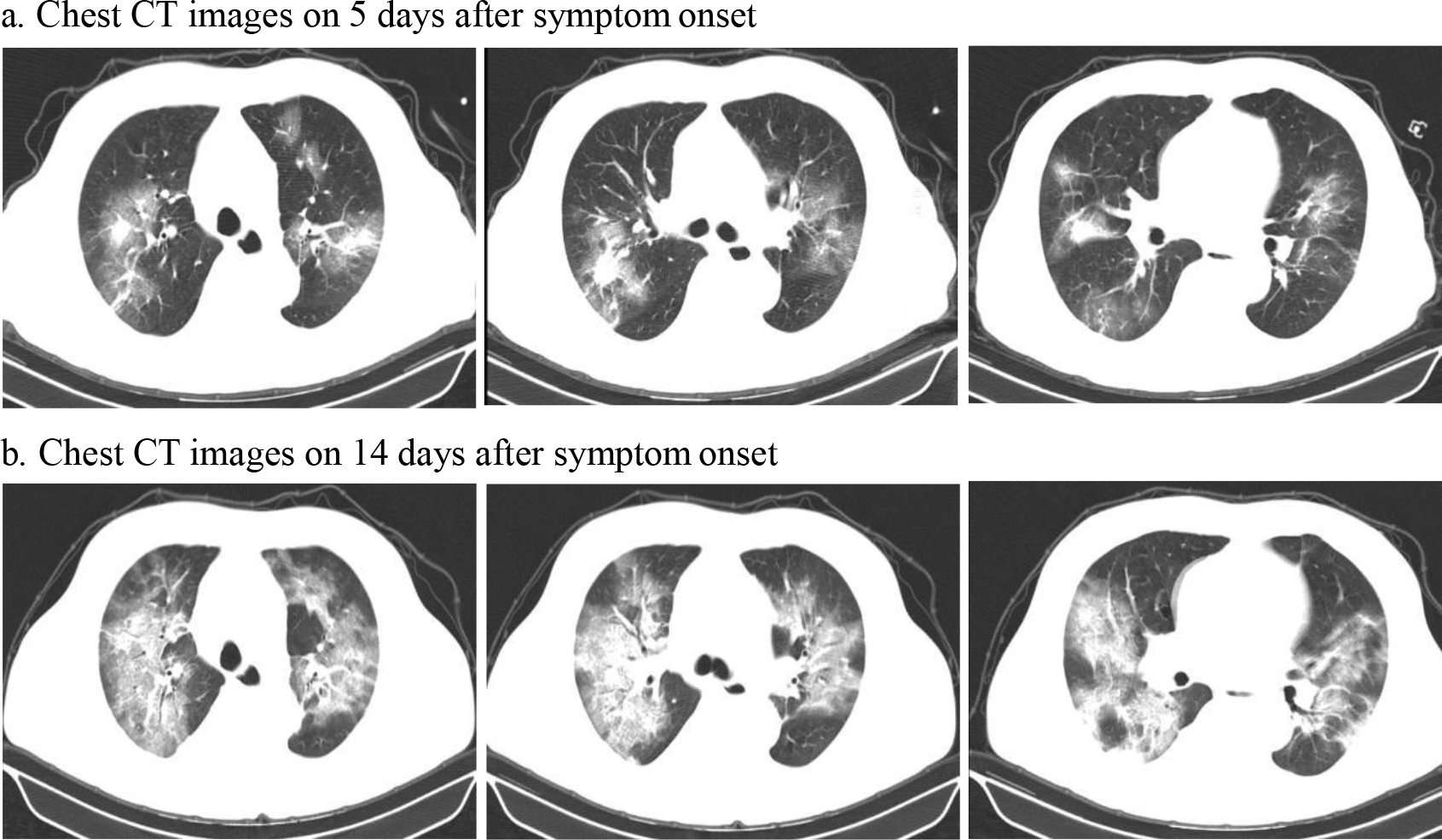
Chest CT images of a 62-year-old patient with COVID-19. a Chest CT images obtained on February 2, 2020, showed ground glass opacity in both lungs on day 5 after symptom onset. b Images taken on February 11,2020, revealed the lesions increased rapidly, and the patient died on February 16,2020.

### Treatment and outcome of adult discharged patients with COVID-19

To avoid to hypoxemia and acute respiratory distress,(ARDS) 147(76.6%) patients received oxygen by nasal catheter, 34(15.3%) patients received oxygen by mask, and 16 (8.1%) patients were treated with invasive ventilation in the ICU. Common complications among the 197 discharged patients included acute liver injury, septic shock, acute myocardial injury, acute renal injury and ARDS. Most patients were treated with antiviral drugs (Oseltamivir), 151(76.6%) patients received antibiotics(moxifloxacin and cefperazone-sulbactam), and several patients were treated with glucocorticoids (75, 38.1%) and immunoglobulin(52, 26.9%). Additionally, 81(41.1%) patients were treated with Chinese traditional medicine. By the end of Febururay 26, 28 (14.2%) patients had died, and 169(85.2%) patient was discharged and transferred to designated hospital for observation. The average of hospital stay was 15.08 days (Table 4).

## Discussion

Coronavirus was a large RNA virus, including SARS-CoV, MERS-CoV, HCoV-OC43, HCoV-229E, HCoV-NL63 and HCoV-HKU1. SARS-CoV and MERS-CoV may induce sever respiratory syndrome, but the remains only caused mild upper respiratory infectious.2019-nCoV was a new subtype, with high incidence and rapid infection, it has spread widely in China and abroad, involving Italy, German, Korean, Iran, and Japan ^[8-9]^. At present, there was no vaccine and antiviral drugs against this disease. Therefore, Extensive studies of clinical features on COVID-19 have not only a better understanding of coronavirus transmission,but have also been key to detect quickly and accurately.

COVID-19 diagnosis was based on exposure histories, symptoms, laboratory tests, chest CT and RT-PCR. In the early phase of the epidemic, almost all patients worked at or lived around the Huanan Seafood Whole Market in Wuhan ^[10]^. With COVID-19 spread rapidly in Hubei province, Wuhan-related exposures was regarded as a key criterion. Previous retrospective research demonstrated that 86% cases had a Wuhan-related exposures history^[11]^. Similarly, our study revealed most of infected patients lived around Xiling strict, because of many residents working at China Gezhouba Group Corporation, returning from Wuhan. The common symptoms of COVID-19 patients were fever, fatigue and cough, some patients may present with abdominal symptoms ^[12]^. While a few patients without any symptoms were detected accidentally by chest CT ^[11]^.Thus, it is difficult to suspect the disease without auxiliary examinations.

The significant of laboratory examinations was blood routine, the results suggested leukocytes were mainly normal or decreased, and lymphocytes reduced evidently in many COVID-19 patients ^[13]^. A recent research indicated that lymphocytes,especially CD3^+^ lymphocytes and CD45^+^ lymphocytes, decreased significantly in comparison with non-COVID patients, while there was no significantly difference in quantities and proportions of neutrophils and monocytes^[14]^.Results of this analysis showed lymphocytes and eostinophils decreased obviously without leukocytes increasing in most patients, and neutrophils,monocytes and platelet mainly kept at normal range,it was consistent with previous reports. In term of chest CT, the typical image features were peripheral, subpleural ground glass opacities in the lower lobes. However, a study of CT manifestations demonstrated that it was approximately 14% COVID-19 patients with atypical CT features ^[15]^. Real time RT-PCR was considered a standard assessment for COVID-19,but the sensitivity was lower than chest CT scans, RT-PCR was 71%, chest CT scans reached 98%^[16]^.Thus, it was essential to perform chest CT for detecting the suspected patients.

As described before, there was currently no clinical approved antiviral drugs and vaccine for coronavirus infections except for supportive treatment ^[17]^. Immunoglobulin may enhance the ability of anti-infection in severely ill patient, and steroids are considered in those patients with ARDS. Oxygen inhalation was important to avoid to ARDS for the COVID-19 patients with hypoxemia. In this study, 76.6% patients received oxygen by nasal catheter, and 90% received antiviral therapy, and 38.1% received glucocorticoids. CDC reported that the case-fatality rate (CFR) of COVID-19 the overall case--fatality rate (CFR) of COVID-19 was 2.3% in confirmed cases, and CFR among critical cases may reach 49.0% ^[11]^. Nevertheless, most of patients were still treated in the hospital. Another retrospective study suggest the CFR was about 11% ^[13]^. Statistical analysis of this study found that the CFR was 14.2%, and most of them were over 70 year-old patients accompanying with chronic illness. In the past one month, numerous effective measures, including blocking cities and roads, closing supermarket, monitoring temperature, building a new hospital and setting up medical assistance team, have been carried out to controlling infection source, preventing transmission and reducing death. Until now, there was no new confirmed patients for 13 days,and hospitalized patients decreased sharply.

In conclusion, this retrospective study, based on clinical data, revealed that COIVD-19 mainly infected in older people with comorbidities, and may cause acute severe respiratory syndrome.Chest CT was key to screen those suspected patients, and lymphocytopenia and eosnophils declining without leukocytes increasing may be regarded as an novel evidence in the dignosis of COVID-19.

## Data Availability

The data used to support the findings of this study are available from the corresponding author upon request

## Acknowledgment

None.

